# EDEPS (Early Detection of ExtraPyramidal Symptoms): supervised machine learning models to detect antipsychotics-induced extrapyramidal hand tremor from a mobile device built-in sensors

**DOI:** 10.1101/2024.12.16.24319069

**Authors:** Adam Wysokiński, Aleksandra Zwierzchowska-Kieszek

**Affiliations:** Medical University of Lodz, Department of Old Age Psychiatry and Psychotic Disorders, Czechosłowacka 8/10, 92-216 Łódź, Poland

**Keywords:** parkinsonism, tremor, antipsychotics, supervised machine learning

## Abstract

**Introduction:** Approximately 30% of patients treated with antipsychotics develops extrapyramidal side effects, among which hand tremor is not only common, but also significantly impacting daily activities. No tool for remote assessment of hand tremor is available.

**Materials and methods:** We collected SAS and AIMS scores and digital recordings of health tremor from healthy and schizophrenia patients on antipsychotics. Next, we created and tested a supervised machine learning models for detecting and measuring severity of antipsychotics-induced hand tremor.

**Results:** We present model details, accuracy measures (R^2^ and RMSE for regressors; log loss, AUC, misclassification, rate, accuracy, sensitivity and specificity for classifiers) and analysis of hand tremor spectral analysis.

**Conclusions:** Our model offers a satisfactory accuracy (0.95 to 1.0) and performance, even if only 10 second data is available. Result of the spectral analysis indicate that the dominating frequency of hand tremor in antipsychotics-induced EPS is approximately 5.0 Hz.

## Introduction

Antipsychotic medications remain the foundation of the pharmacological management of schizophrenia, bipolar disorder, psychotic depression and other mental disorders. While having relatively good efficacy, antipsychotics are also associated with extrapyramidal symptoms (EPS), also called drug-induced movement disorders. These include parkinsonism, akathisia and tardive dyskinesia. Recent data indicate that EPS are present in approximately one-third of all subjects treated with antipsychotics, posing a serious burden and distress to these patients [1].

Anticholinergic medications (e.g. trihexyphenidyl and procyclidine) are very often use to reduce antipsychotic-induced EPS. While they may be effective in that regard, they pose several serious adverse consequences. Due to their mechanisms, they may induce blurry vision, constipation, memory impairment. Also, they do not reduce the risk of development of tardive dyskinesia. Finally, even when used in therapeutic doses, anticholinergics may cause recurrence or exacerbation of psychosis [2]. Therefore, the recommended approach to EPS is to avoid medications that may cause these side effects and switch the treatment as soon as they are detected.

Several scales are used to screen for and evaluate extrapyramidal symptoms, including Simpson-Angus Scale (SAS) [3], Abnormal Involuntary Movement Scale (AIMS) [4], Extrapyramidal Symptom Rating Scale (ESRS) and its abbreviated version (ESRS-A) [5]. All the scale available today require physical contact with the patient and cannot be performed remotely. During the recent COVID-19 pandemic many psychiatrists who were forced to switch to telepsychiatry and while it exhibited considerable success in its effectiveness [6], in many clinical situations (such as evaluating extrapyramidal symptoms) there remains a significant unfilled demand.

The objective of this study is to address the issue of limited availability of tools for evaluating extrapyramidal symptoms remotely. We have attempted to develop a computer model allowing prediction of extrapyramidal hand tremor presence and severity using easily available data from a mobile device (phone or tablet) built-in gyroscopic sensors.

## Material and Methods

This research consisted of two parts. First, we collected recording data from healthy subjects and schizophrenia patients with extra-pyramidal hand tremor to train and validate the regressor and classifier models. Next, we have collected a separate group of healthy subjects and schizophrenia patients with extra-pyramidal hand tremor to validate the models accuracy.

All study subjects were adults, aged 18-65 years and gave informed consent for participating in the study. The study protocol had been approved by the local Bioethics Committee. Inclusion criteria for the no tremor group (NTRM) were: no diagnosis of Axis I mental disorder (current or past), no diagnosis of neurological disorders (current or past), no hand tremor (assessed by the study personnel on the study enrollment). Inclusion criteria for the tremor (TRM) group were: diagnosis of schizophrenia, treatment with any type of antipsychotic(s) for at least 6 weeks prior the study enrollment, no treatment with anti-Parkinson’s medications (levodopa, carbidopa, pramipexole, rotigotine, ropinirole, amantadine, anticholinergics or MAO-B inhibitors), and the presence of hand tremor (assessed by the study personnel on the study enrollment). From the TRM subjects we have collected SAS and AIMS (items 8, 9 and 10 only) scores.

All data were recorded using MATLAB Mobile and Apple iPad (2018 model). Sampling rate was set to 50.0 Hz, data were recorded for acceleration, orientation and angular velocity (X, Y, Z axes). During the recording the device was held horizontally in an outstretched hand. In NTRM group the right hand was used for recording. In the TRM group the hand with more severe tremor was used. After recording was was completed, data was exported as the MATLAB .MAT file and processed locally. During the processing each recording the first second of the recording is removed and the remaining signal is spliced into 10-second epochs. As we collected at least 1 minute of recording from each subject, the processing produced at least 5 to 6 epochs for each subject. Resulting data was saved as .CSV files for further analysis.

### Computer model

The EDEPS (**E**arly **D**etection of **E**xtra**P**yramidal **S**ymptoms) modeling toolbox is licensed under the 2-Clause BSD License. EDEPS.jl source code and all training / testing data are available in the toolbox repository at https://codeberg.org/AdamWysokinski/EDEPS.jl. The machine learning algorithms were implemented using Random Forrest regressor and classifier based on Julia 1.10 [7]. In the modeling stage we have tested several different algorithms (including K-Nearest Neighbours (KNNs), Naive Bayes, Bagged Trees, Adaptive Boosting, AdaBag, Support Vector Machines (SVMs) with various kernels, Logit Boost, NNET) to find that the Random Forrest gave best results both as regressor and classifier.

The training process consists of several stages. Imported raw data (acceleration and angular velocity in X, Y, Z axes – 6 signals in total; we rejected orientation data since they were not improving the accuracy) are imported and converted into Power Spectral Density (PSD) data, calculated for each signal. PSDs are calculated using the Welch’s periodogram and Hanning window. These data are further transformed using Z-Score scaler and split randomly into training and validating groups (70:30 ratio). Final model is retrained on the complete dataset. The classifier is a supervised, Random Forrest model with the following parameters: minimum samples split = 2, minimum purity increase = 0.0, n trees = 250, sampling fraction = 0.99, feature importance = impurity). The regressor is a supervised, Random Forrest model with the following parameters: minimum samples split = 2, minimum purity increase = 0.0, n trees = 260, sampling fraction = 0.99, feature importance = impurity). The regressor is trained using the following scores: SAS score (0 to 3), AIMS item 8 score (0 to 3), AIMS item 9 score (0 to 3), AIMS item 10 score (0 to 3) and sum of AIMS 8, 9 and 10 scores score (0 to 9). Parameters of both models were established iteratively based on misclassification rate (for classifier) and root mean squared error (RMSE) (for regressor) of observed vs. predicted data. We regularly re-test the model using new subjects. If the prediction parameters (misclassification rate and RMSE) of the re-test subjects worsen, we add new subjects data into the dataset and repeat the models’ parameters optimization and retrain the models.

In the prediction process imported raw data are processed as described above and first entered into the regressor mode, to predict SAS, AIMS 8, 9, 10 and total AIMS scores. Next, the data is entered into the classifier model to predict whether the subject has tremor or not. Additionally, we calculate adjusted classification prediction. The adjustment procedure is based on weighting the classification probability using weights based on predicted SAS or AIMS 8 and AIMS 9 scores. These weights were established experimentally based on prediction accuracy measurements (−0.1 for score < 1, +0.1 for score in [1, 2), +0.2 for score in [2, 3) and +0.3 for score ≥ 3).

The prediction algorithm can be run locally (see README.md file in the EDEPS.jl repository for instructions).

## Results

### Training and validation evaluations

For the classifier model the training data included 2338 10-second epochs (NTRM: 1323 (56.6%), TRM: 1015 (43.4%)). For the regressor model the training data included 1053 10-second epochs. Regressor model training and validation accuracy measures are shown in Table 1. The distribution of the final model errors (measured value - predicted value) for predicted SAS and AIMS scores is shown in Figure 1. Classifier model training and validation accuracy measures are shown in Table 2.

**Table 1.** Regressor model accuracy measures (training).

| **model** | **RMSE** | **R<sup>2</sup>** |
| --- | --- | --- |
| regressor: SAS training | 0.21 | 0.96 |
| regressor: AIMS 8 training | 0.17 | 0.94 |
| regressor: AIMS 9 training | 0.22 | 0.91 |
| regressor: AIMS 10 training | 0.28 | 0.92 |
| regressor: AIMS training | 0.66 | 0.90 |
| regressor: SAS validation | 0.62 | 0.65 |
| regressor: AIMS 8 validation | 0.39 | 0.65 |
| regressor: AIMS 9 validation | 0.61 | 0.32 |
| regressor: AIMS 10 validation | 0.80 | 0.41 |
| regressor: AIMS validation | 1.57 | 0.47 |
| regressor: SAS final model | 0.24 | 0.95 |
| regressor: AIMS 8 final model | 0.21 | 0.90 |
| regressor: AIMS 9 final model | 0.30 | 0.85 |
| regressor: AIMS 10 final model | 0.36 | 0.87 |
| regressor: AIMS final model | 0.85 | 0.84 |

**RMSE:** root mean squared error

**Table 2.** Classifier model accuracy measures (training).

| **model** | **log**<br>**loss** | **AUC** | **misclassification**<br>**rate** | **accuracy** | **sensitivity**<br>**(TPR)** | **specificity**<br>**(TNR)** |
| --- | --- | --- | --- | --- | --- | --- |
| classifier: training | 0.04 | 1.0 | 0.0 | 1.0 | 1.0 | 1.0 |
| classifier: validation | 0.14 | 0.99 | 0.06 | 0.94 | 0.95 | 0.93 |
| classifier: final model | 0.04 | 1.0 | 0.0 | 1.0 | 1.0 | 1.0 |

**AUC:** area under curve; **TPR:** true positive ratio; **TNR:** true negative ratio

**Figure 1.**
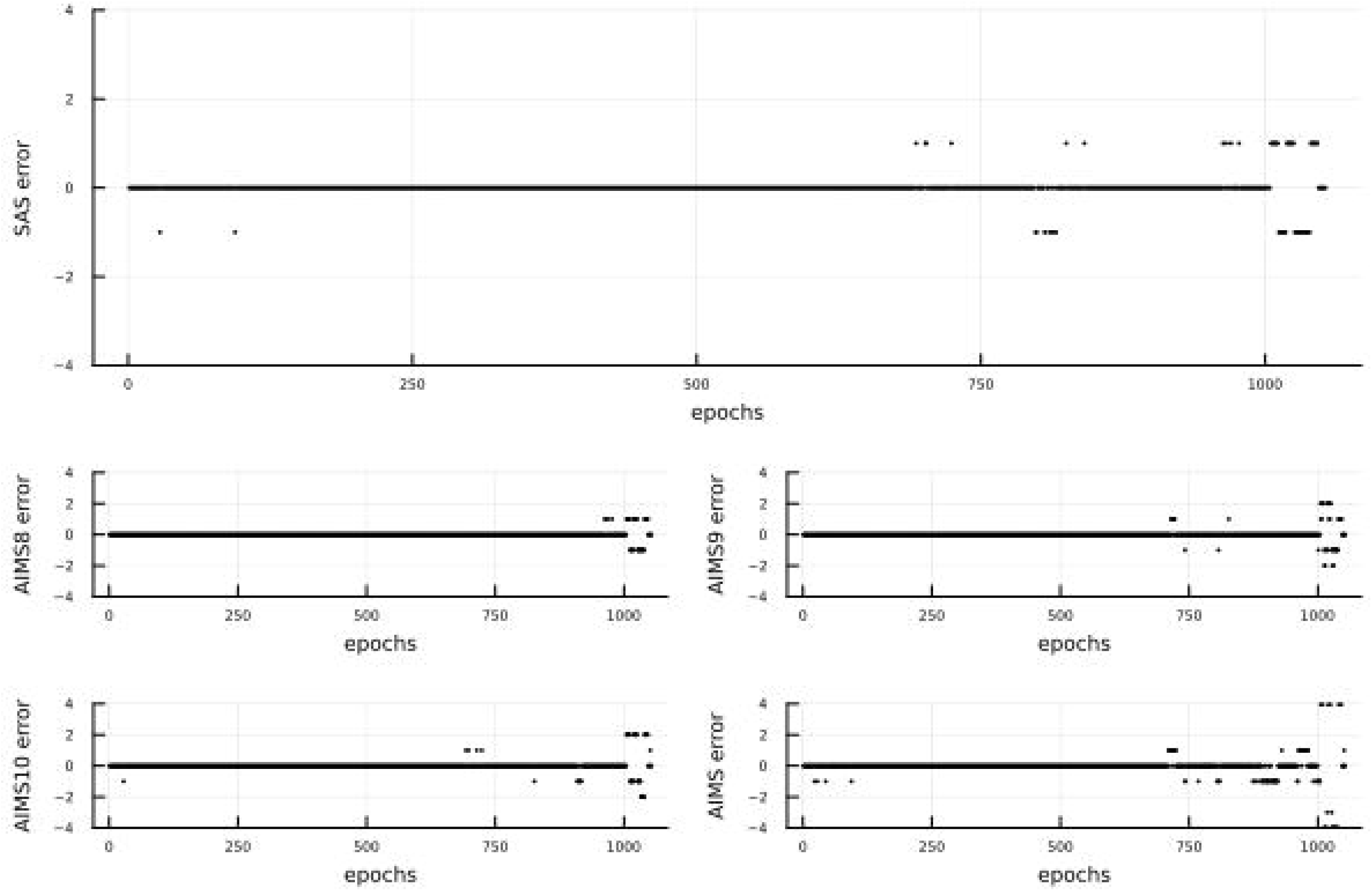
Prediction error for the final regressor model (training). error = measured value - predicted value

### Testing evaluations

For the regressor model the testing data included 204 epochs (29 subjects), while for the classifier model the testing data included 613 epochs (88 subjects, NTRM: n=45 (51.1%) TRM: n=43 (48.9%)). In the testing group we have performed two types of predictions. First, we ran regression and classifier models using all available epochs for each subject. Next, we did the same but using only the first 10-second epoch. Regressor model accuracy measures are shown in Table 3, while the distribution of the errors (measured value - predicted value) for predicted SAS and AIMS scores is shown in Figure 2. Classifier model accuracy is shown in Table 4.

**Table 3.**
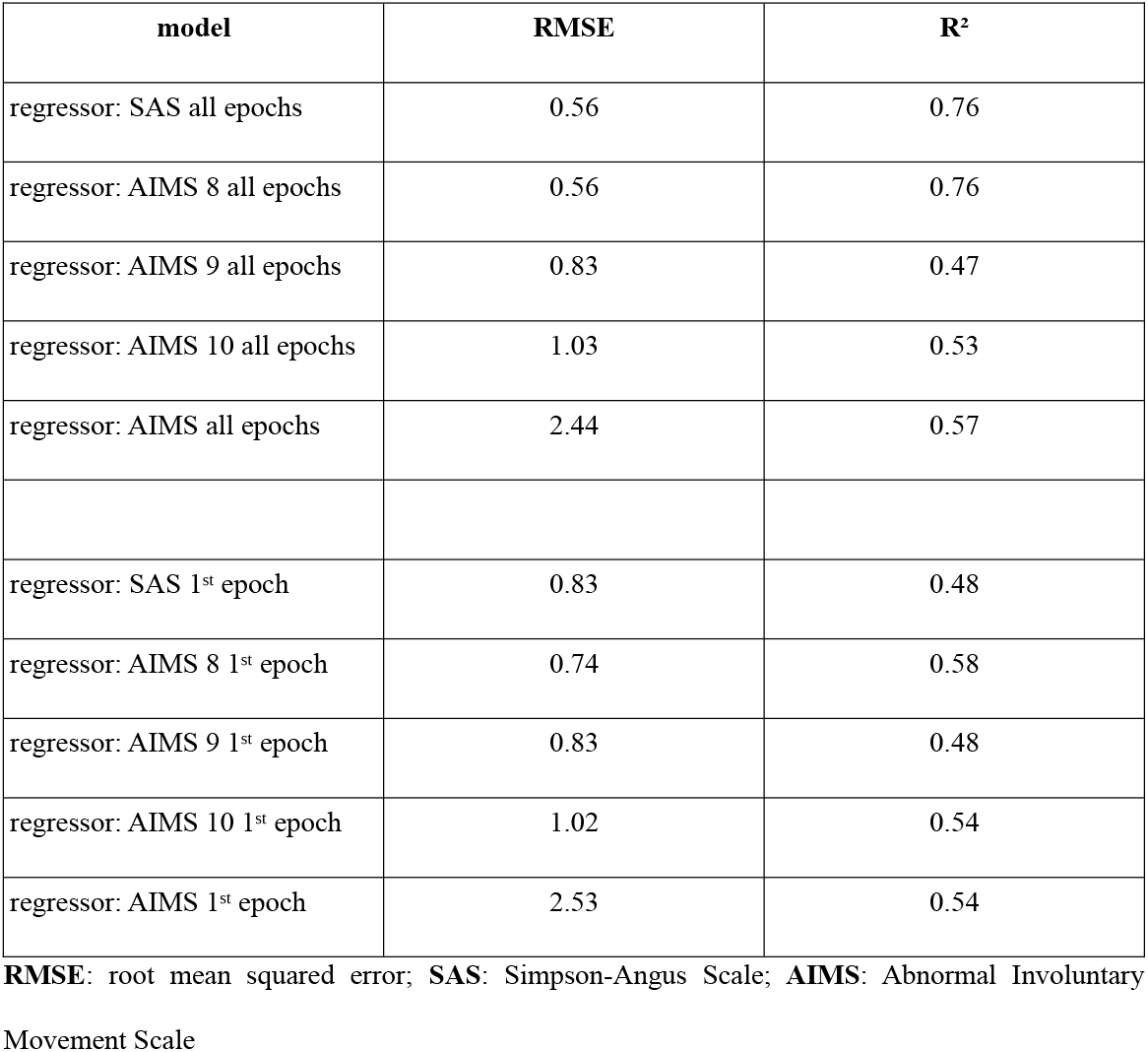
Regressor model accuracy measures (testing).

| **model** | **RMSE** | **R<sup>2</sup>** |
| --- | --- | --- |
| regressor: SAS all epochs | 0.56 | 0.76 |
| regressor: AIMS 8 all epochs | 0.56 | 0.76 |
| regressor: AIMS 9 all epochs | 0.83 | 0.47 |
| regressor: AIMS 10 all epochs | 1.03 | 0.53 |
| regressor: AIMS all epochs | 2.44 | 0.57 |
| regressor: SAS 1<sup>st</sup> epoch | 0.83 | 0.48 |
| regressor: AIMS 8 1<sup>st</sup> epoch | 0.74 | 0.58 |
| regressor: AIMS 9 1<sup>st</sup> epoch | 0.83 | 0.48 |
| regressor: AIMS 10 1<sup>st</sup> epoch | 1.02 | 0.54 |
| regressor: AIMS 1<sup>st</sup> epoch | 2.53 | 0.54 |

**RMSE:** root mean squared error; **SAS:** Simpson-Angus Scale; **AIMS:** Abnormal Involuntary Movement Scale

**Table 4.** Classifier model accuracy measures (testing).

| **model** | **log**<br>**loss** | **AUC** | **misclassification**<br>**rate** | **accuracy** | **sensitivity**<br>**(TPR)** | **specificity**<br>**(TNR)** |
| --- | --- | --- | --- | --- | --- | --- |
| classifier: all epochs | 0.15 | 0.99 | 0.05 | 0.95 | 0.98 | 0.93 |
| classifier: 1<sup>st</sup> epoch | 0.16 | 0.99 | 0.05 | 0.95 | 0.91 | 1.0 |
| classifier: all epochs<br>(adjusted) | NA | NA | 0.0 | 1.0 | 1.0 | 1.0 |
| classifier: 1<sup>st</sup> epoch<br>(adjusted) | NA | NA | 0.0 | 1.0 | 1.0 | 1.0 |

**NA:** not available
**AUC:** area under curve; **TPR:** true positive ratio; **TNR:** true negative ratio

**Figure 2.**
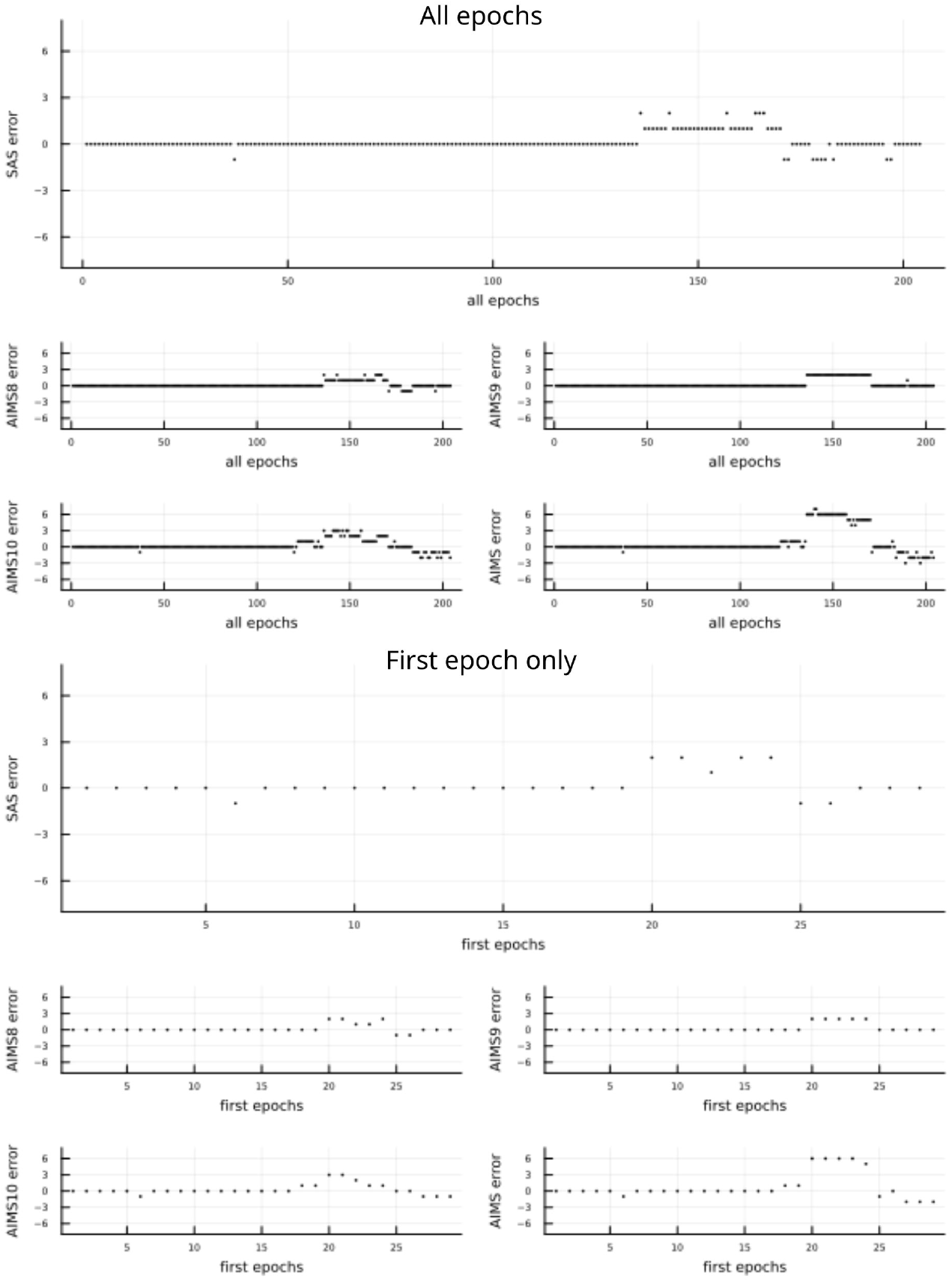
Prediction error for the final regressor model (testing). error = measured value - predicted value

### Power spectrum density

Averaged power spectrum density was calculated for both groups, separately for each signal channel. There is a distinctive peak in the power distribution at around 5 Hz for all analyzed channels (acceleration and angular velocity) in patients with hand tremor, see Figure 3.

**Figure 3.**
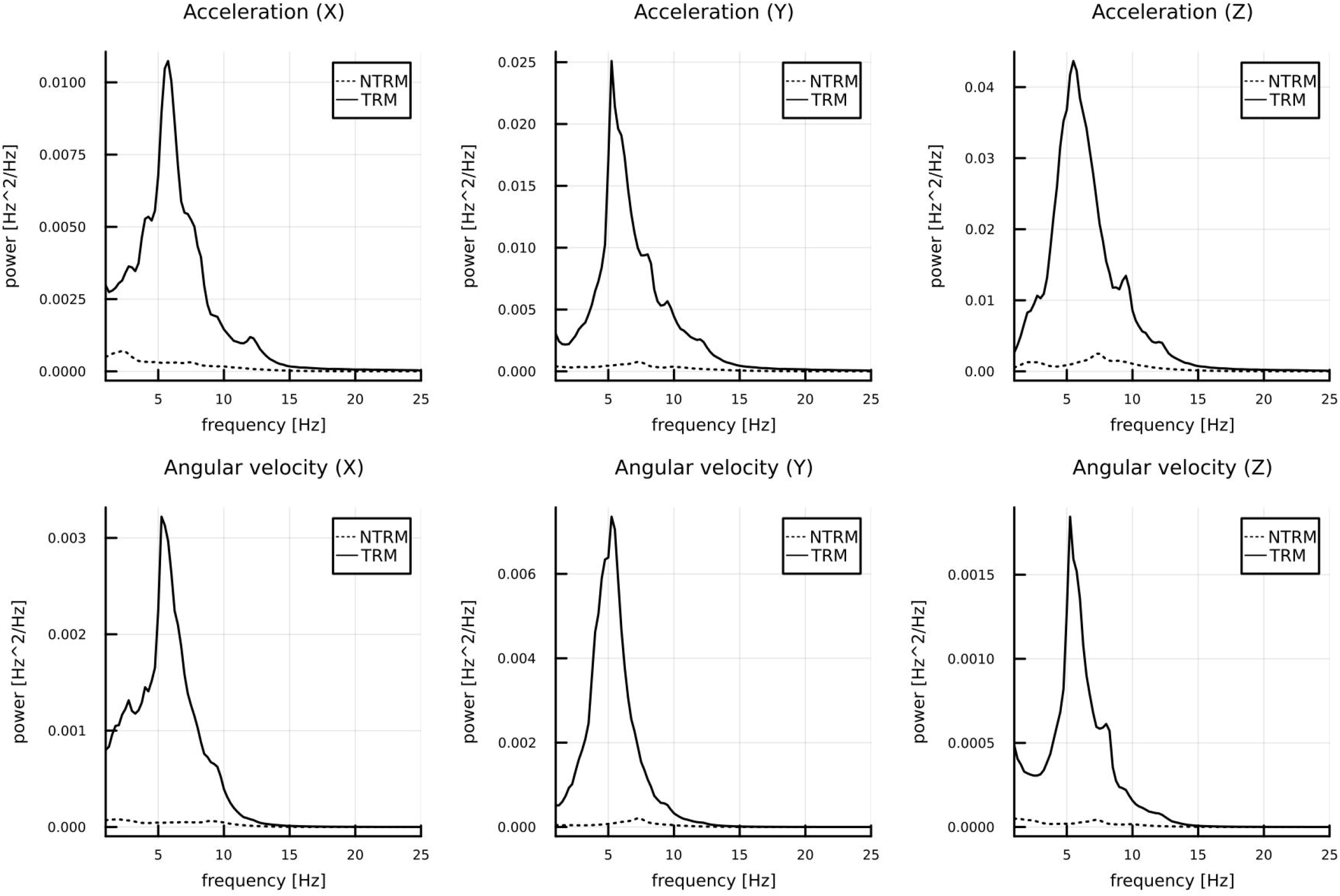
Averaged power density plots. **NTRM**: no tremor subjects **TRM**: tremor subjects

## Discussion

To our knowledge this is the first model that allows predicting extrapyramidal hand tremor in schizophrenia patients using mobile device built-in gyrosensors. Using the model we were able to detect which patients have tremor and accurately predict their severity using SAS and AIMS scores. The model offers a satisfactory accuracy and performance, even if only 10 second data is available. Result of the spectral analysis indicate that the dominating frequency of hand tremor in antipsychotics-induced EPS is approximately 5.0 Hz.

Sun et al. published a study in which a wearable wrist sensor was used to assess the Parkinson’s disease (PD) action tremor [8]. They have tested various supervised algorithms and found that Support Vector Machines (SVMs) performed the best with over 90% F1 scores. Automated tremor detection in PD patients using smartwatch sensor data was analyzed by [9]. They stated that smartwatches can capture tremors in patients with PD. However, tremors may not always manifest during smartwatch-based data capture, ultimately affecting the reproducibility. The authors suggest that smartwatch integration would further enable improved longitudinal assessment at home. In 2021 Fuch et al. demonstrated that tremor data gathered using the smartphones was useful for the constructing of machine learning models that can be used to monitor patients with essential tremor. In this study the mobile phone was strapped to the wrists of patients with essential tremor. That procedure requires skilled assistance of medical personnel and therefore its application for schizophrenia patients, who often suffer from cognitive deterioration, might be problematic.

There are many task-based tools for measuring fine motor skills that may be applied in patients with hand tremor, extensive list is discussed by Norman and Héroux [10], examples are finger tapping, rotating a coin, drawing a spiral. While they may serve a screening purpose, quantitative interpretation of their results may be limited.

A very interesting approach was tested by Crespo et al. These authors explored the use of a digitizing tablet to assess motor symptoms in schizophrenia or bipolar disorder patients [11]. It was possible to measure the task quantitatively (e.g. entropy of velocity and pressure). Also, such test, when performed using a regular tablet, would be performed remotely. It would however require a special digital pen (to record screen pressure), so again its practical application would be limited. There was an attempt to quantify bradykinesia during clinical finger taps using a gyrosensor in patients with Parkinson’s disease [12]. This approach works well, but requires an external gyrosensor attached to the finger and therefore would have a limited applicability in daily practice. As in every model, there are certain limitations and imperfections. We did not compare patients with other types of hand tremor (e.g. essential tremor, lithium-induced or related to alcohol withdrawal syndrome). Patients with such commodities would be excluded from the study, therefore we assume that our model is able to differentiate between various types of hand tremor. Also, it’s quantitative performance is not perfect, especially for AIMS scores. We did not perform test-retest reliability tests. Since the Random Forrest models are pretty complex for larger number of trees (here 260 for regression and 250 for classification), interpretation of their weights is difficult.

## Data Availability

All data produced in the present study are available upon reasonable request to the authors

## Acknowledgments

None

